# Efficacy of a self-management program using an eHealth system to reduce symptom severity in patients with irritable bowel syndrome simultaneously with changes in gut microbiota: a randomized controlled trial

**DOI:** 10.1101/2022.12.22.22283873

**Authors:** Jun Tayama, Toyohiro Hamaguchi, Kohei Koizumi, Ryodai Yamamura, Ryo Okubo, Jun-ichiro Kawahara, Kenji Inoue, Atsushi Takeoka, Antonius Schneider, Shin Fukudo

## Abstract

**Objective:** This study aimed to evaluate whether a self-management program using the eHealth system could reduce symptom severity in patients with irritable bowel syndrome (IBS). Impact of the intervention on quality of life and gut microbiota were also examined.

**Design:** This study was designed as an open label, simple randomized controlled trial comparing an intervention group that attended an eHealth self-management program and a treatment as usual group. Participants were Japanese women between the ages of 18 and 36. Forty symptomatic IBS individuals who met the inclusion criteria were recruited and randomly assigned to the two groups. The eHealth group received 8 weeks of unlimited access to the self-management program containing a wide variety of e-learning content. Participants’ severity of IBS symptoms, the main outcome, was assessed using the irritable bowel syndrome-severity index (IBS-SI) at baseline and 8 weeks. The secondary outcomes of participants’ quality of life and gut bacteria were also assessed at baseline and week 8.

**Results:** There was a significant difference in the net change in IBS severity index (IBS-SI) score between the eHealth and treatment as usual group (−50.1; 95% CI, −87.6 to -12.6; p = 0.010). The eHealth group had significantly lower IBS-SI scores following 8 weeks of intervention compared with the baseline scores (t = − 3.2, p < 0.01). The implementation of the eHealth program was accompanied by improvement of quality of life and decrease of phylum-level Cyanobacteria occupancy, respectively.

**Conclusion:** The implementation of eHealth for IBS was shown to reduce IBS symptoms.

**Key Messages:**

- **What is already known on this topic –** eHealth programs based on diet and probiotic use have shown good results in reducing IBS symptoms
- **What this study adds** – a self-management program with an e-learning component based on a successful self-help guidebook for IBS
- **How this study might affect research, practice or policy** – The proposed eHealth model reduces symptoms and improves the quality of life of IBS patients, providing an efficient and cost-effective intervention option to be adopted in policy and practice, and creates scope for future research in food intake, exercise, and sleep management through eHealth for IBS.

## INTRODUCTION

Irritable bowel syndrome (IBS) is a functional gastrointestinal disorder characterized by marked abnormality in brain-gut interaction in the absence of major organic abnormality [1]. The main pathophysiological features of IBS are dysmotility of the lower gastrointestinal tract [2], visceral hypersensitivity [3], and psychological abnormalities [4]. IBS is highly prevalent worldwide, with an adult prevalence rate of 10.1% with Rome III and 4.1% with Rome IV [5]. In Japan, the prevalence is as high as about 9.3% with Rome III and 2.2% with Rome IV [5]. IBS is also associated with impaired daily functioning, and a pronounced decline in quality of life [6]. For example, issues such as interference with daily activities, health-related anxiety, food avoidance, sexual concerns, and poor relationships, can arise [7]. Although IBS is not life-threatening, it is very damaging in socioeconomic terms [8, 9]. In prior research, the magnitude of the economic impact of having IBS has been estimated to be 1.1 to 6.0 times greater than that of non-IBS controls [10].

The clinical practice guidelines of American College of Gastroenterology and Japanese Society of Gastroenterology for the management of IBS recommend non-pharmacologic therapies along with pharmacotherapy as treatment for IBS [11, 12]. In both human [8, 13, 14] and animal studies [15], gastrointestinal symptoms of IBS or IBS-like gastrointestinal function have been found to be exacerbated by worsening of psychiatric symptoms or corresponding behavior, which can improved by psychological recovery. Therefore, psychotherapy as well as drug therapy is essential for improving IBS symptoms. Psychotherapy using hypnosis [16, 17, 18], and more recently, non-hypnotic approaches such as cognitive-behavioral therapy [19, 20], mindfulness [21, 22], and psychoeducation [23, 24] contribute to the improvement of IBS symptoms. Other non-pharmacologic therapies such as exercise therapy [25, 26] are also known to be effective. With speed at which the relationship between the intestinal microbiota and IBS symptoms is being elucidated in recent years [27], dietary therapies to reduce IBS symptoms by improving the intestinal environment, such as the low FODMAP (fermentable oligosaccharides, disaccharides and monosaccharides and polyols) diet [27, 28], are also attracting attention.

Non-pharmacological treatment of IBS often incorporates self-management methods in which patients actively control their own symptoms. Self-management of IBS can ameliorate disease and economic burdens [29, 30]. For example, through cognitive-behavioral therapy [19, 20], patients can increase their sense of control over IBS symptoms by setting their own goals and working on their own tasks. The low FODMAP diet controls IBS symptoms by helping patients learn how to eat and manage their food in an appropriate manner [27, 28]. Self-management of IBS using a self-help guidebook [31, 32] is a non-pharmacological intervention that requires a significant amount of patient effort [30, 31, 32]. A randomized controlled trial using a self-help guidebook with 420 patients with IBS in the United Kingdom found that the intervention group had 60% fewer visits to primary care, less severe IBS symptoms, and 40% lower annual cost per patient compared to the control group one year after the intervention [30]. A prospective observational study of 71 IBS patients in Germany using a self-help guidebook reported a significant increase in quality of life after six months of intervention [32]. Thus, although using self-help guidebooks requires a great deal of effort from IBS patients, it contributes to improvement in symptoms and quality of life, and reduction in medical costs.

The present study introduces eHealth, a web-based practice that assists healthcare providers in ambulatory care, and evaluates its potential to enhance self-management in IBS patients. The application of eHealth for IBS treatment and follow-up has been shown to alleviate symptoms, optimize patient compliance, improve quality of life, and reduce economic burden [33, 34, 35, 36]. The advantage of eHealth is that it facilitates IBS patients to engage in individualized self-management therapy [33]. Many eHealth programs provide both a mechanism for managing patient data, such as symptoms, and for patients to access IBS-related content via e-learning at their convenience [34, 36, 37]. A previous study of 34 patients with IBS examined the effect of eHealth based probiotic treatments and a low FODMAP diet on reducing IBS symptoms and found comparable symptom reduction with both treatments [34]. Regarding the IBS self-help guidebook [30, 31, 32] mentioned earlier, its content is yet to be validated in eHealth form.

This study’s primary objective was to verify the hypothesis that an eHealth-based self-management program can reduce the severity of IBS symptoms.

## PATIENTS AND METHODS

### Study design

This study was an open label, simple randomized controlled trial with an intervention group receiving a self-management program through eHealth and a treatment as usual (TAU) group. This study was registered at https://center6.umin.ac.jp/cgi-open-bin/ctr/ctr_view.cgi?recptno=R000047461 (UMIN Clinical Trials Registry, UMIN000042552).

### Patient and public involvement

Patients were not involved in the design, implementation, reporting, or dissemination plans of our research.

### Participants

Participants comprised 40 symptomatic IBS patients who met the ROME ? criteria and were enrolled from university students in Japan. Previous studies have shown that women and younger people are at higher risk of IBS [5], based on which we set the inclusion criteria as Japanese women within the age range of 18 to 36 years. The exclusion criteria were having been previously treated with medication for IBS, having any preexisting psychiatric disorders, and other organic gastrointestinal diseases. All patients provided written informed consent to participate in this study. The study protocol was approved by the Ethics Committee of Saitama Prefectural University (no. 20048) and the study was conducted in accordance with the ethical standards of the Declaration of Helsinki, 1964 and its later amendments at the 64th World Medical Assembly, Fontareza, 2013. Compliance with the study protocol was verified by the access logs of the eHealth system.

### The eHealth program

For our eHealth program, we modified the content of the existing self-help guidebook for IBS, which includes six chapters: “Personal experiences of IBS,” “Understanding IBS,” “What you can do to help yourself,” “More ways to manage your IBS,” “Medical treatments,” and “Summary and sources of information” [31, 32]. The five chapters in our program also included content from previous studies[31, 32] (Table 1). The eHealth program for IBS was designed to support learning not only on computers but also on mobile devices. The program also made it possible to download content and store it locally on personal devices. Hence, participants could access content and learn at a time and place of their choosing. Each chapter comprised text in an e-book format and narrated video, for increased accessibility and usability [38]. Quizzes were placed at the end of each chapter. The content was available for viewing for a period of eight weeks and could be downloaded and viewed, such that the participants could access and learn as much as they wanted at their own schedule and convenience. The goal was to study each chapter at least once and complete the quiz at the end. Assessments were conducted before the start of the eHealth intervention (baseline) and at the end of the intervention (at 8 weeks).

**Table 1.**
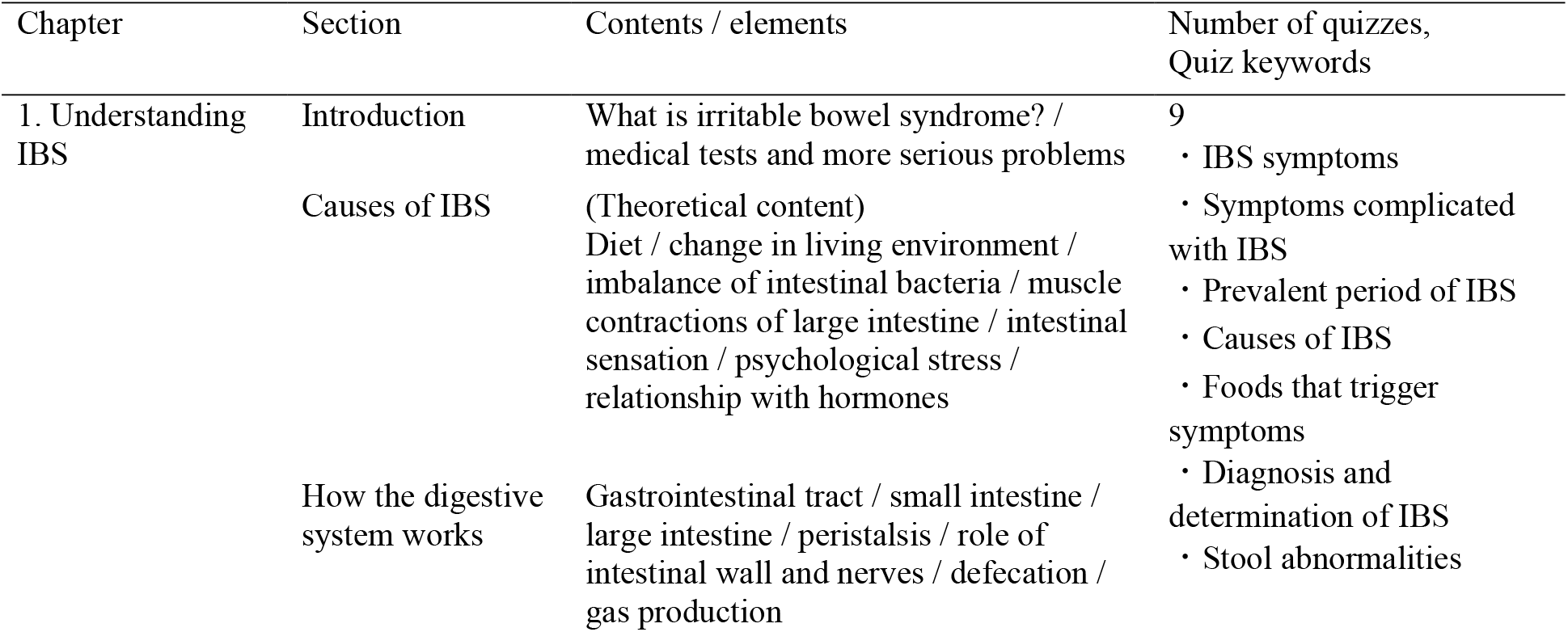
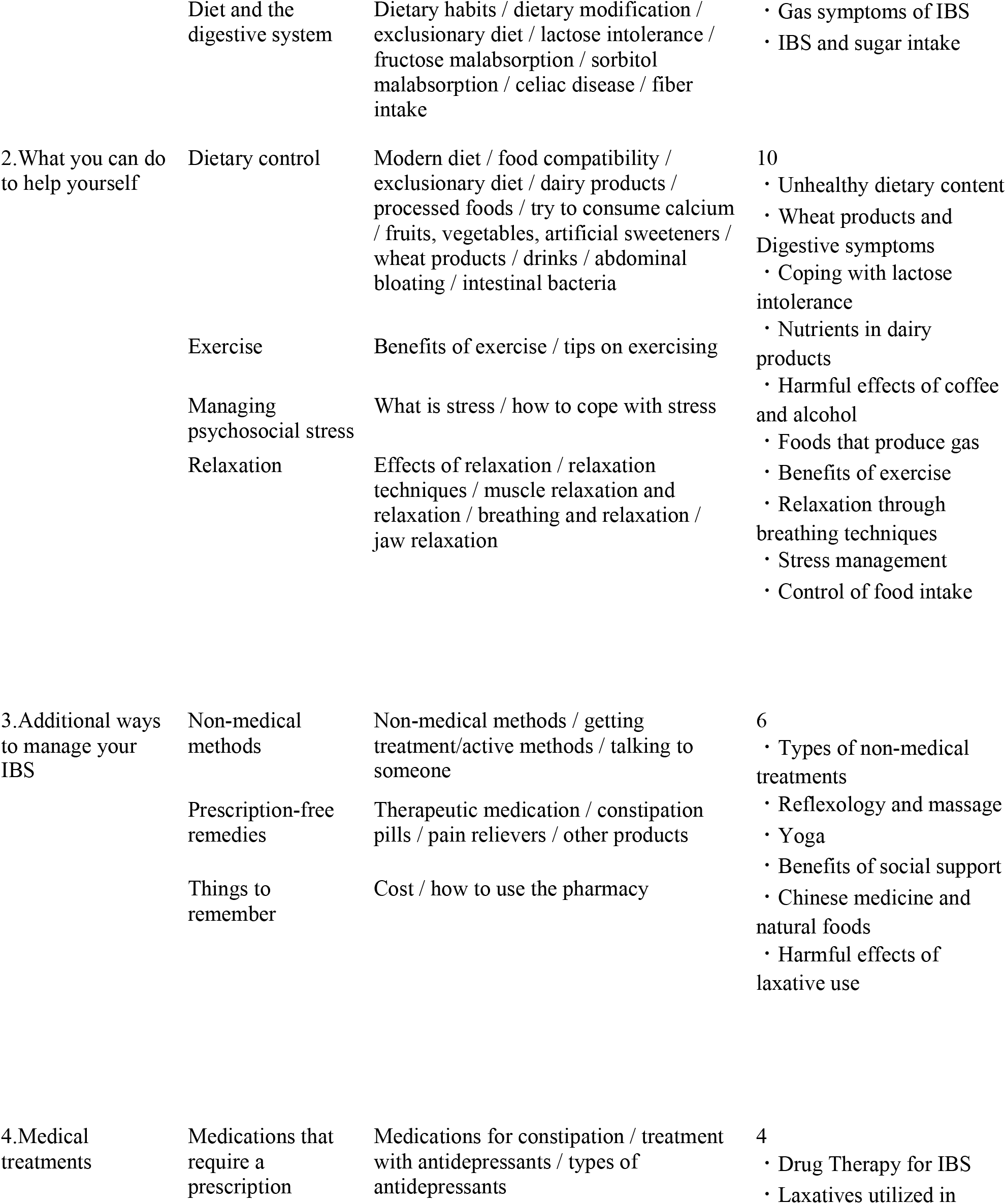
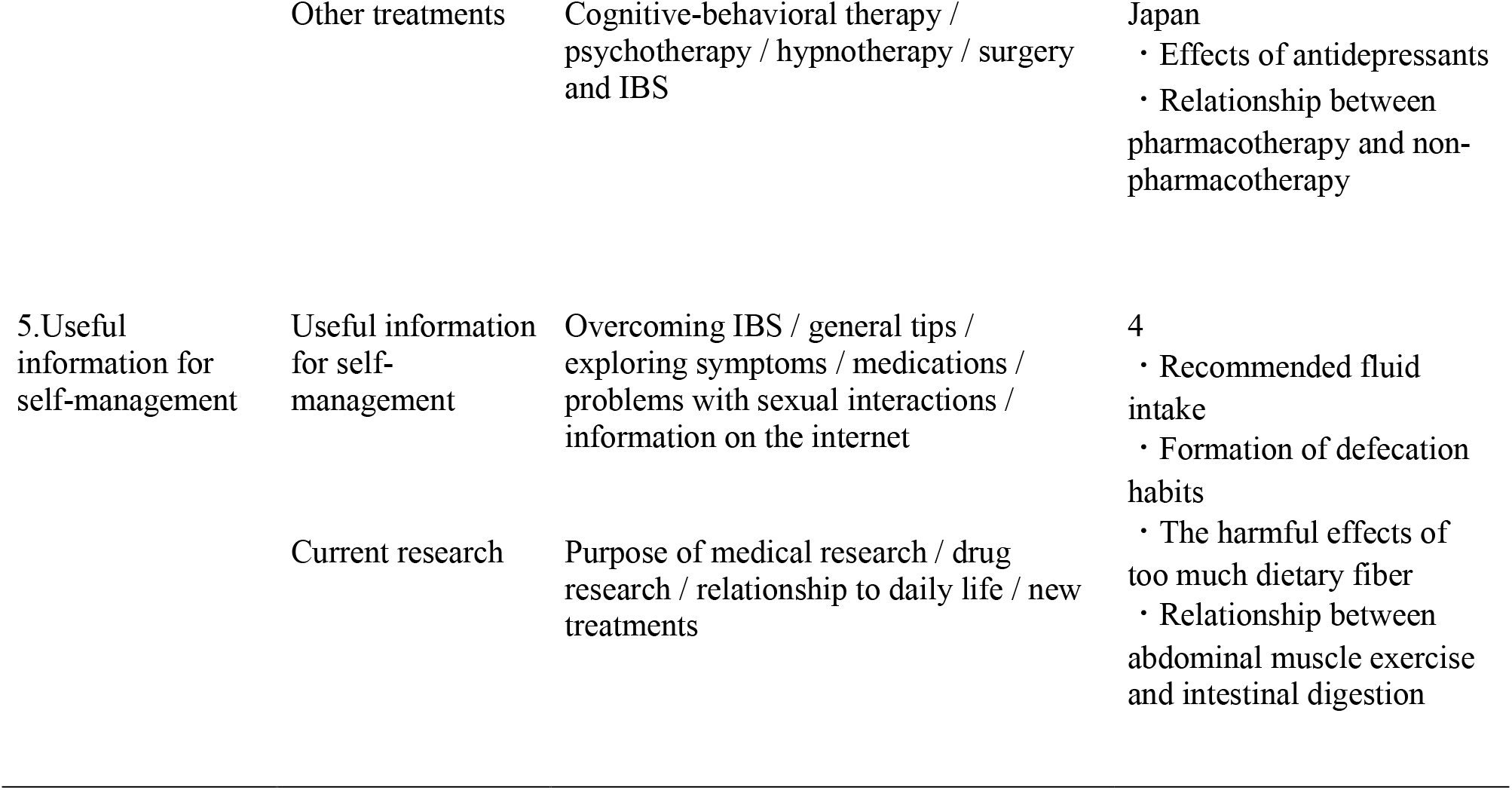
Elements included from each chapter of the eHealth program

### Treatment as usual (TAU)

For the TAU group, access to the eHealth program was not implemented; however, assessments similar to the intervention group were conducted at the same time points (baseline, 8 weeks). After the study was conducted, the participants were given access to the eHealth program and were allowed to access the same content as the eHealth group for a period of approximately 8 weeks. The TAU group faced no restrictions on treatment of any kind, including drug therapy.

### Randomization

The participants were randomly and equally assigned to the eHealth intervention or TAU groups using a random number table created with Microsoft Excel by trial statisticians. Because all participants and investigators could be aware of which group each participant was assigned to, the design of this study corresponded to open label.

### Outcomes

The primary outcome measured was the total score on the IBS severity index (IBS-SI, IBS-symptom severity scale, IBS-SSS) [39, 40] at 8 weeks after the intervention. Secondary outcomes were the total score on the IBS-quality of life measure (IBS-QOL) [7, 41], compositions of the gut microbiota (at the phylum, order, class, family, and genus levels), and α-diversity indices of the gut microbiota. In addition, the intake based on the low FODMAP diet was qualitatively evaluated.

### Sample Size

The sample size was determined based on a previous study [27] that investigated the improvement effect of IBS-SI by a 3-week non-pharmacological treatment for patients with IBS. The study found a reduction in mean IBS-SI score following the intervention (Mean ± S.D. treatment group; IBS-SI score 208.0 ± 74.8 and TAU group 290.0 ± 106.0). From this we estimated that ≥ 17 individuals per group were required for a difference in IBS-SI score ≥ 82.0 (S.D. = 31.2) with an α level of 0.05 (2-tailed) and 80% power.

### Bacterial DNA extraction and microbiome analysis

Bacterial DNA was extracted from feces samples using a nucleic acid extraction system PI-1200 (Kurabo, Osaka, Japan). Each library was prepared according to the Illumina 16S Metagenomic Sequencing Library Preparation Guide with a primer set 27Fmod/338R targeting the V1–V2 region of 16S rRNA genes. The 251-bp paired-end sequencing of the amplicons was performed on a MiSeq system (Illumina, CA, USA) using a MiSeq Reagent v2 500 cycle kit. All steps from the trimming of the paired-end reads FASTQ files which were obtained via 16S rRNA amplicon sequencing to the gut microbiota analysis, were performed using QIIME 2 [42]. First, the raw sequence results were demultiplexed, and the DADA2 algorithm was used to identify microbial operational taxonomic units. We then classified operational taxonomic units into five taxonomic rank categories (phylum, order, class, family, and genus) using the SILVA 132 reference database at 99% similarity. The Shannon index (H’) and Simpson index (1-D), which are a measure of α-diversity, were calculated using the following equations at the genus level: H’ = -∑*pi*ln*pi* and D = ∑*pi*^2^, where *pi* is the relative abundance (%) of *genus i* in the community.

### Statistical analysis

Data were expressed as mean ± S.D. Analysis of covariance (ANCOVA) was used to assess the differences between mean scores, 95% confidence interval values, and p-values for each outcome. Covariates for the ANCOVA were the continuous variables of age and body mass index (BMI), the discrete variable of IBS subtype (IBS with diarrhea (IBS-D), IBS with constipation (IBS-C), mixed IBS (IBS-M), and unsubtyped IBS (IBS-U)) and the continuous baseline scores for each outcome. A two-tailed test was used with the α level set at 0.05%. The p value was calculated using Bonferroni correction. In accordance with a previous report [40], we defined an IBS-SI score of 175 or higher as moderate to severe IBS, and calculated the percentage of IBS for each time course to test the difference in proportions. Furthermore, we applied the linear discriminant analysis effect size (LEfSe) [43] with default settings to determine the features of the gut microbiota (at the phylum, order, class, family, and genus levels), that can likely explain the differences in each group (eHealth vs. TAU).

## RESULTS

### Demographic data

Prospective participants (n = 160) received a recruitment packet approved by the ethics committee and consented to share their contact information with the research team. Of the 160 approached participants, 99 who were non-IBS at screening were excluded, resulting in 61 potential participants assessed for eligibility. Of the 61, 21 withdrew from participation. Finally, of the 40 remaining patients, 21 were randomly assigned to the eHealth group and 19 to the TAU group. Subsequently, all 40 participants (100%) completed the four weeks intervention program, and the four weeks follow-up. All 21 participants (100%) in the eHealth group accessed the content of all five chapters present in the eHealth program and each of the chapter quizzes at least once (Figure 1).

**Figure 1.**
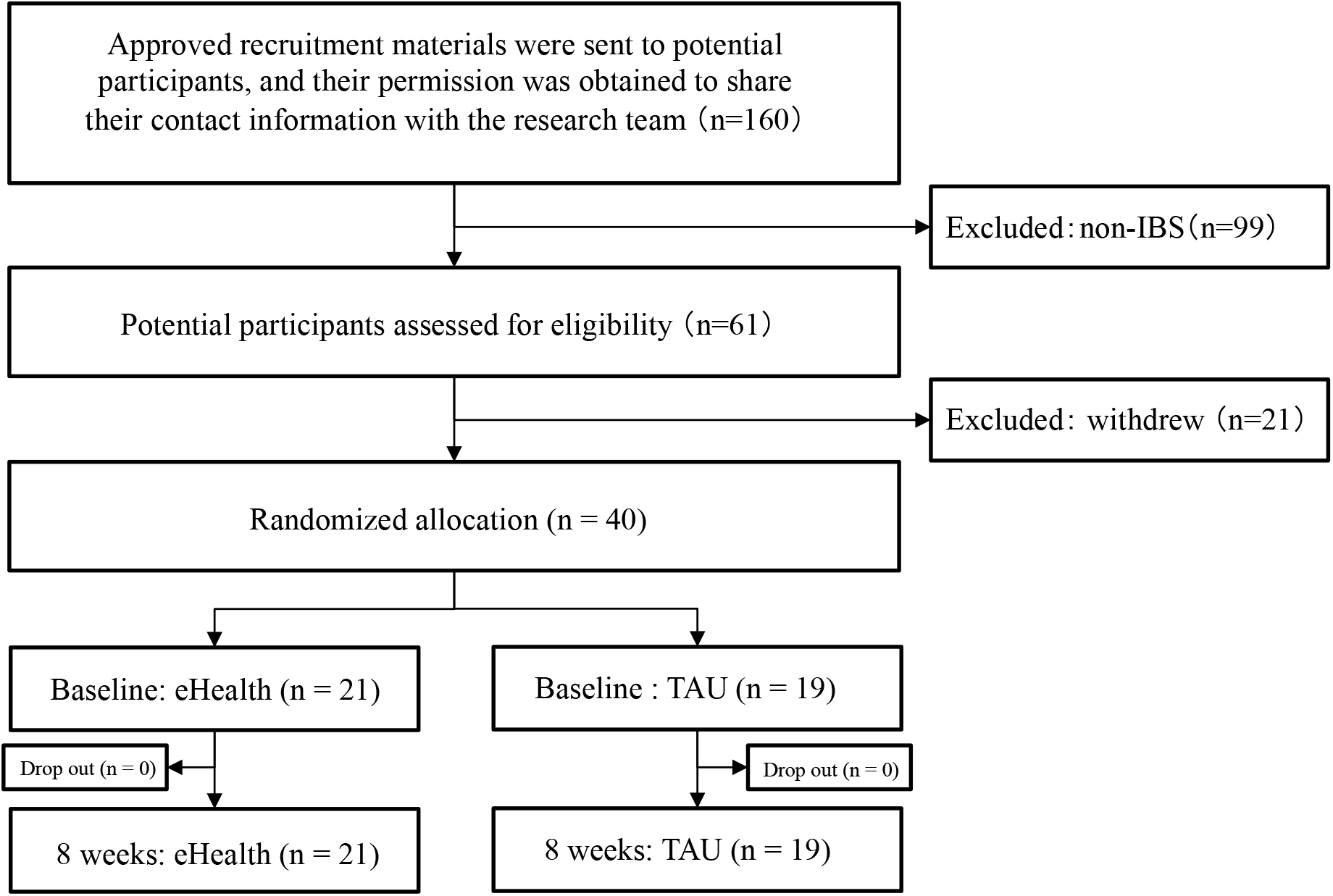
Recruitment, eligibility, and randomization of participants in this study. Of the 160 patients who agreed to participate in the study, 99 were non-IBS at screening and 21 later withdrew. Of the 40 IBS symptomatic individuals, 19 were randomly assigned to the eHealth group for an 8-week eHealth intervention and 21 to the TAU group.

Table 2 represents the baseline demographic data. Patients were well matched for age, BMI, and IBS subtype between groups (Table 2). The total scores of IBS-SI and IBS-QOL at baseline, two of the outcomes measured in this study, were also well matched. No significant differences were observed between the two groups.

**Table 2.**
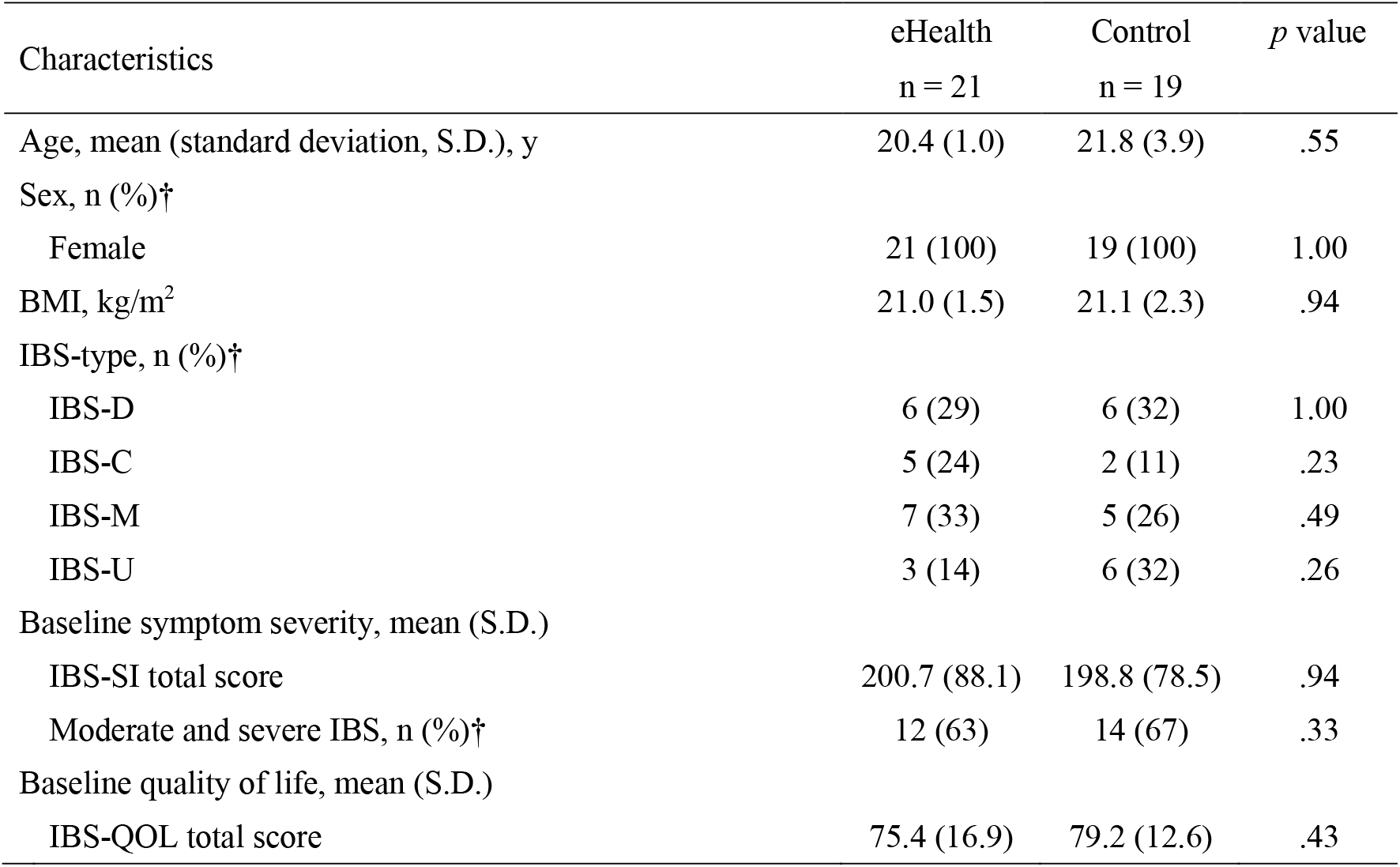
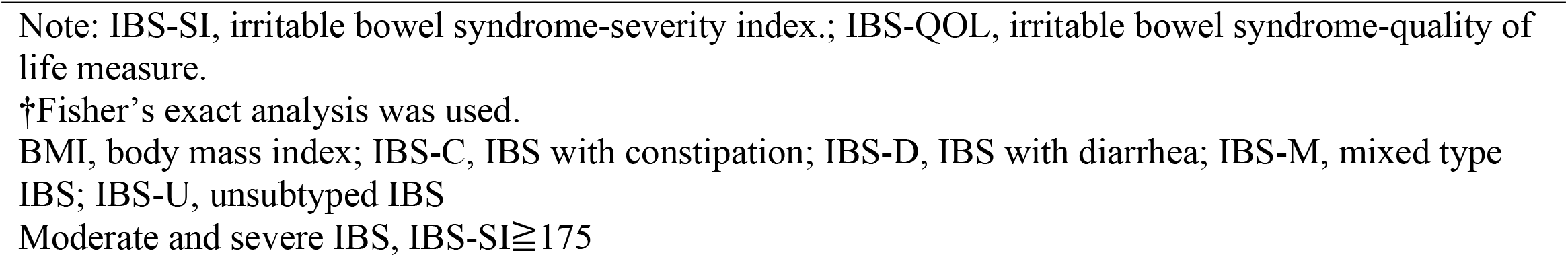
Baseline characteristics of participants.

### Primary outcome measure – IBS-SI

Table 3 summarizes the data at baseline and 8 weeks for the primary outcome, the IBS-SI score. There was a significant difference in the net change in IBS-SI scores between the eHealth and TAU groups (−50.1; 95% CI, −87.6 to -12.6; p = 0.010). Furthermore, the eHealth group had significantly lower IBS-SI scores following 8 weeks of treatment when compared with their baseline scores (t = − 3.2, p < 0.01). Figure 2 shows a time course plot of the change in total IBS-SI scores in the eHealth and TAU groups.

**Table 3.** IBS-SI score at baseline and 8 weeks.

| Group | n | Baseline<br>Mean (S.D.) | 8 weeks<br>Mean (S.D.) | Paired *t*-test |  | Net change<br>(95% CI) | ANCOVA<br>*p* value |
| --- | --- | --- | --- | --- | --- | --- | --- |
|  |  |  |  | *t* | *p* value |  |  |
| eHealth | 21 | 200.7 (88.1) | 131.9 (55.3) | -3.2 | < 0.01 | -50.1 (-87.6, -12.6) | 0.010 |
| Control | 19 | 198.8 (78.5) | 205.9 (77.5) | 0.5 | < 0.61 |  |  |

Note: TAU, treatment as usual; IBS-SI, irritable bowel syndrome-severity index. ANCOVA adjusted for age (continuous variable), BMI (continuous variable), IBS subtype (IBS-C, IBS-D, IBS-M, IBS-U), and baseline IBS-SI score (continuous variable).

**Figure 2.**
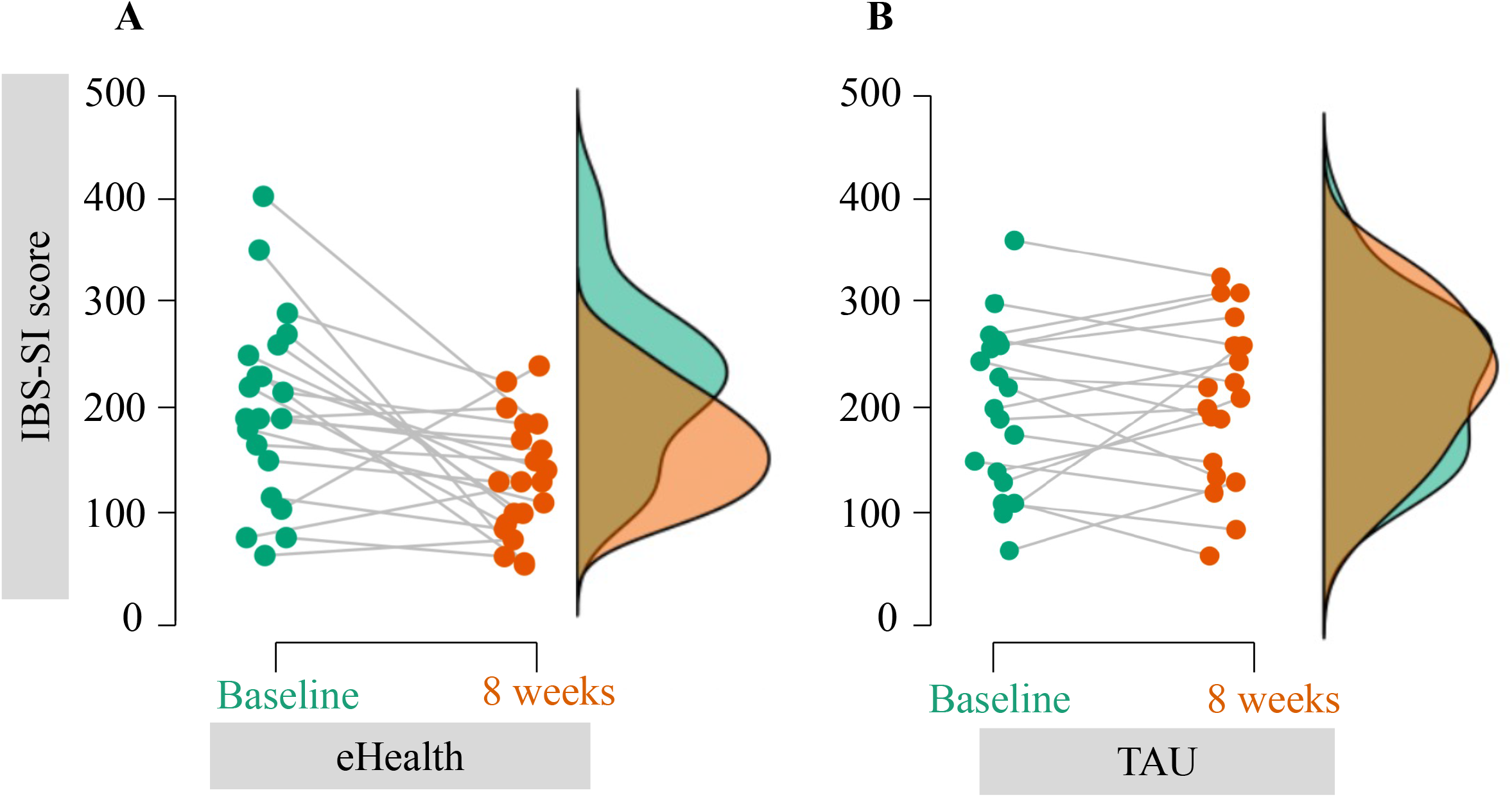
Time course plots of changes in the total score of IBS-SI in the eHealth and TAU groups. TAU means treatment as usual. A: Plots of the eHealth group. B: Plots of the TAU group. The vertical axis represents the total score of IBS-SI. Colored clouds on the right panel show the total score of IBS-SI distributions according to survey periods; green = baseline, and orange = 8 weeks. The ocher areas in the cloud plot are areas where the two colors are transparent and overlap.

### Secondary outcome measure – IBS-QOL

Table 4 summarizes the data for the secondary outcome, IBS-QOL scores, at baseline and 8 weeks. There was a significant difference in the net change in IBS-QOL scores between the eHealth and TAU groups (6.9; 95% CI, 0.5 to 13.2; p = 0.034). Furthermore, the eHealth group had significantly higher IBS-QOL scores following 8 weeks of treatment when compared with their baseline scores (t = 3.9, p < 0.01).

**Table 4.** IBS-QOL score at baseline and 8 weeks.

| Group | n | Baseline<br>Mean (S.D.) | 8 weeks<br>Mean (S.D.) | Paired *t*-test |  | Net change<br>(95% CI) | ANCOVA<br>*p* value |
| --- | --- | --- | --- | --- | --- | --- | --- |
|  |  |  |  | *t* | *p* value |  |  |
| eHealth | 21 | 75.4 (16.9) | 88.1 (10.8) | 3.9 | < 0.01 | 6.9 (0.5, 13.2) | 0.034 |
| Control | 19 | 79.2 (12.6) | 77.5 (13.8) | -0.5 | < 0.59 |  |  |

Note: TAU, treatment as usual; IBS-QOL, Irritable bowel syndrome-quality of life. ANCOVA adjusted for age (continuous variables), BMI (continuous variables), IBS subtype (IBS-C, IBS-D, IBS-M, IBS-U), and baseline IBS-QOL score (continuous variables).

### Secondary outcome measure – percentage of moderate and severe IBS

Figure 3 shows the time course changes in the percentage of moderate and severe IBS (IBS-SI ≧ 175) in both groups. The percentage of patients with moderate and severe IBS in the time course did not change significantly in the TAU group (63% (n = 12) to 68% (n = 13), χ^2^ = 0.117, p = 0.7323). The percentage of the eHealth group over time was significantly different (67% (n = 14) to 24% (n = 5), χ^2^ = 7.785, p = 0.0053).

**Figure 3.**
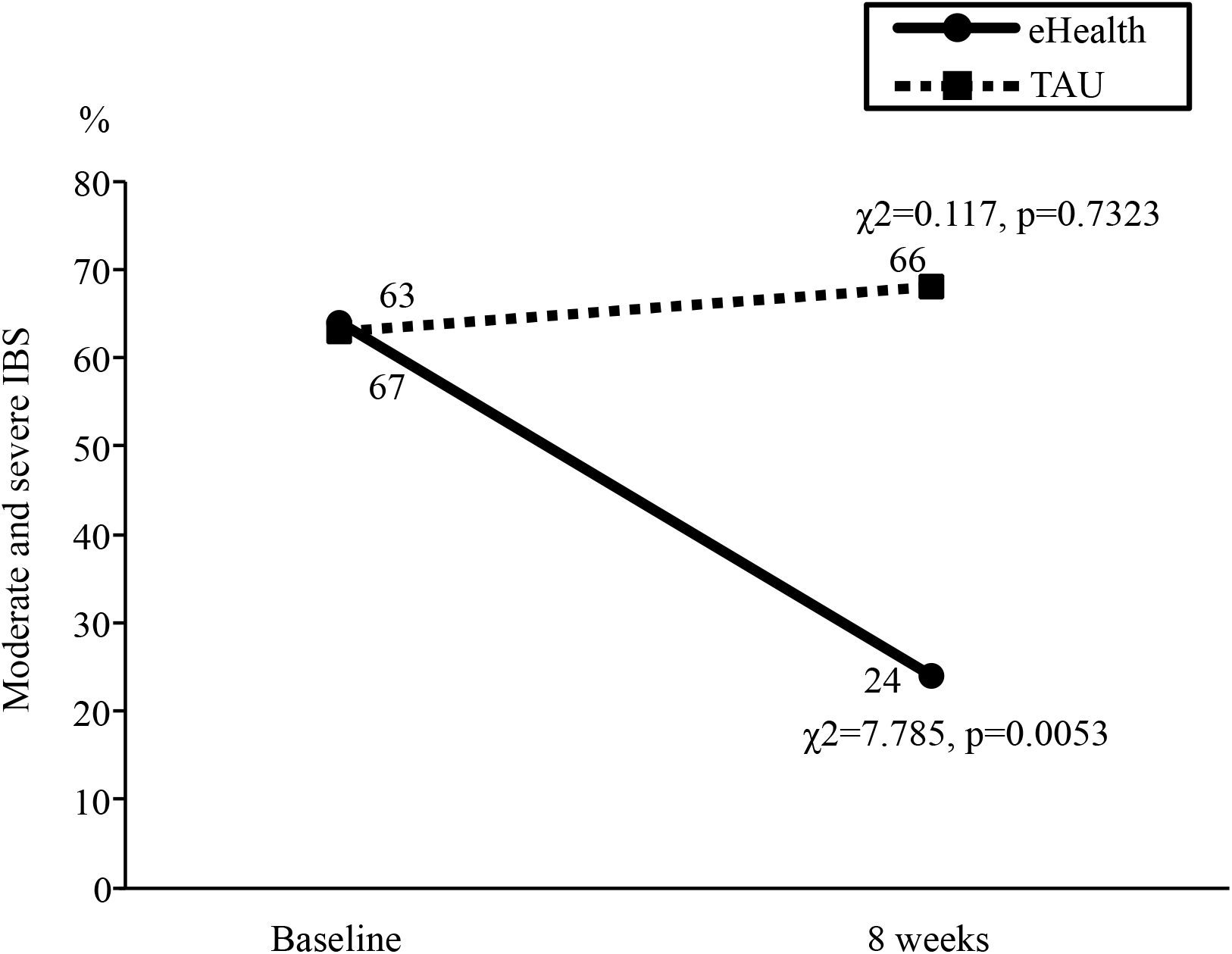
Change in percentage of moderate and severe IBS in both groups. TAU means treatment as usual. The solid line with circled markers is the eHealth group. The dashed line in the square marker is the TAU group.

### Secondary outcome measure – phylum-level compositions and α-diversity indices of the gut microbiota

Table 5 shows the phylum-level compositions and α-diversity indices of the gut microbiota. In phylum-level compositions, there was a significant difference in the net change in phylum Cyanobacteria between the eHealth and TAU groups (−0.01; 95% CI, -0.02 to -0.01; p = 0.001). Otherwise, there was no significant difference in net change between the eHealth and TAU groups in any of the other phylum-level compositions. Furthermore, there was no significant difference in the net change between the eHealth and TAU groups in the α-diversity indices (Shannon and Simpson indices) of the gut microbiota.

**Table 5.**
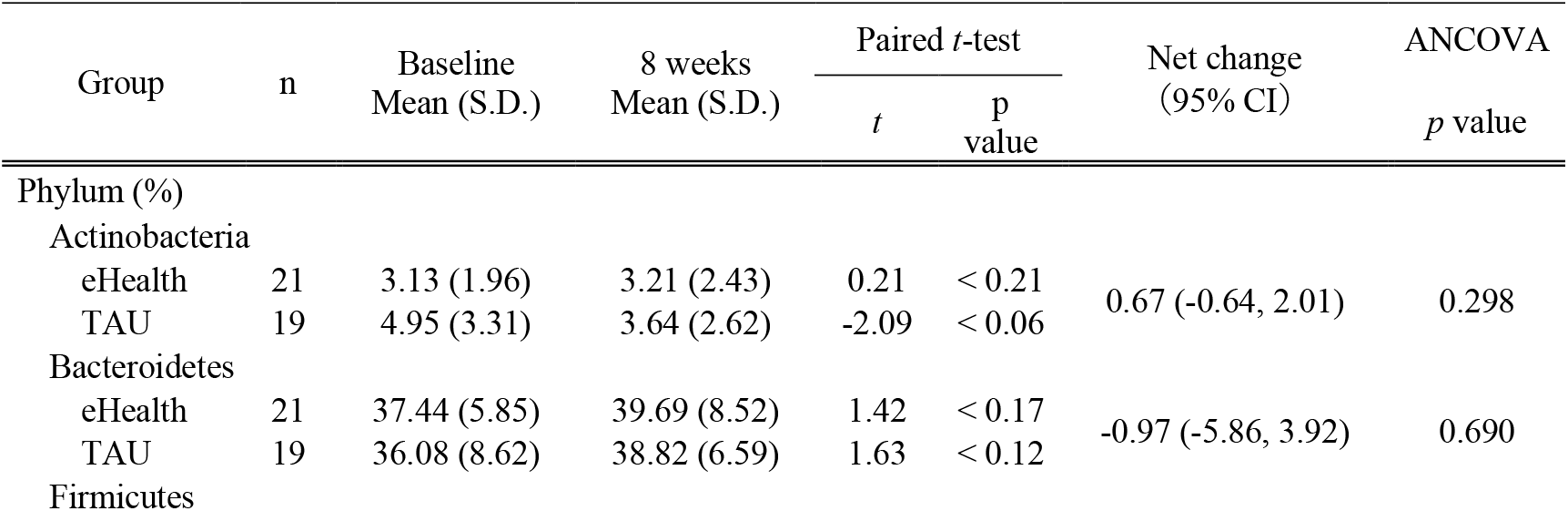
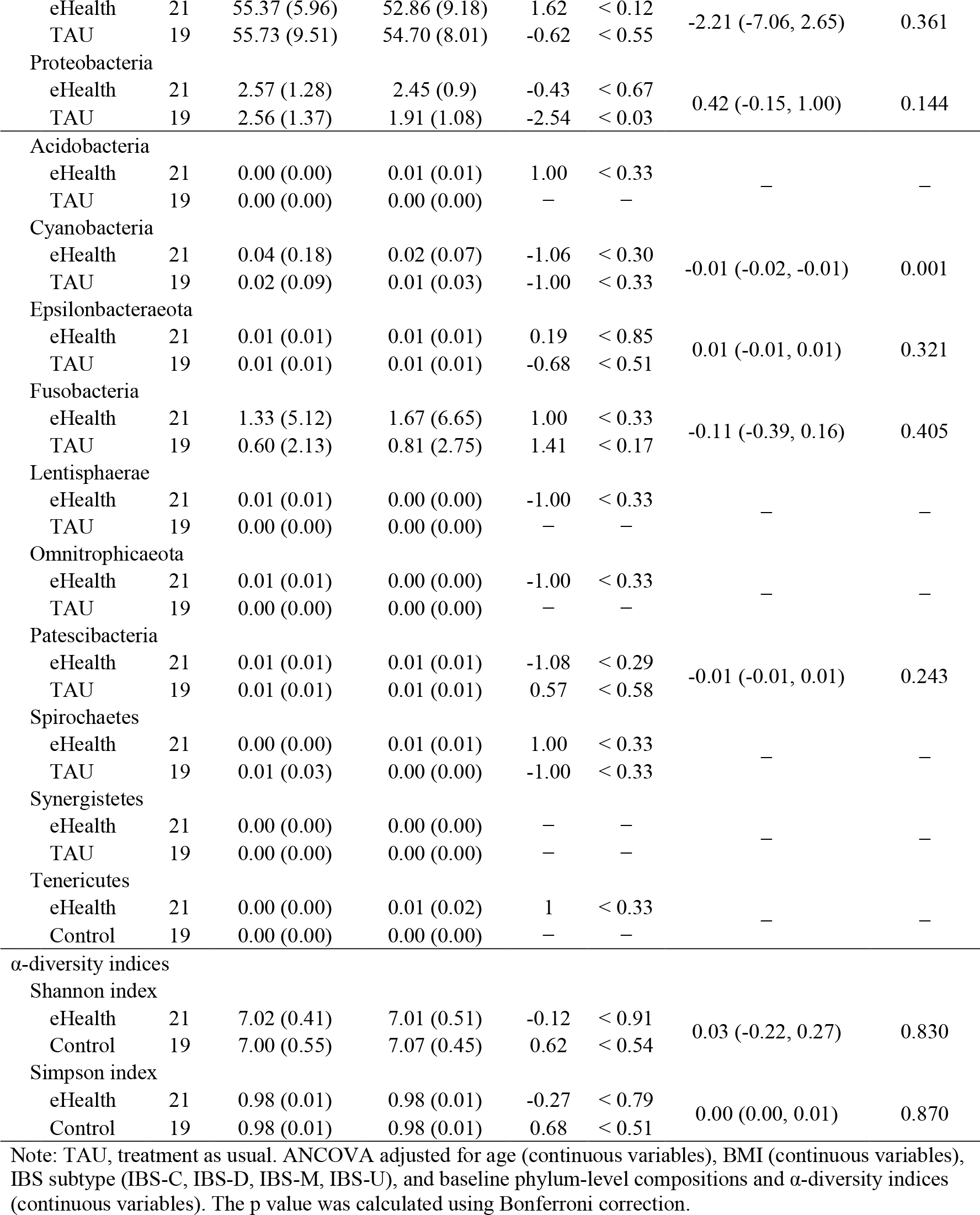
Phylum-level compositions and α-diversity indices of the gut microbiota at baseline and 8 weeks.

### Secondary outcome measure – LEfSe to determine the features of the gut microbiota

We searched for differences in the gut microbiota between the eHealth and TAU groups at each timepoint before and after the intervention by LEfSe, but found no differences (data not shown).

### Qualitative assessment of low FODMAP food intake status

We asked participants the quantity of low FODMAP foods they consumed in the past month from seven food groups: breads and cereals, vegetables, fruit, milk and dairy, protein, nuts and seeds, and beverages. At baseline, there were no group differences in the percentage of the seven food groups consumed. However, at week 8, only milk and dairy products had a higher percentage intake in the eHealth group than in the TAU group (24% (n = 5) vs. 0% (n = 0), p = 0.0230). Regarding the change in time course, there was no significant change in the TAU group for all seven food groups. In the eHealth group, the percentage of those eating low FODMAP foods increased from 71% (n = 15) to 95% (n = 20) in the nut and seed food group (χ^2^ = 4.286, p = 0.0384). Among the eHealth participants, though there were increases of 71% (n =15) to 86% (n =18) in the breads and cereals food group (χ^2^ = 1.273, p = 0.2593), these were not significant. Similarly, in the milk and dairy food group, intake increased from 10% (n = 2) to 24% (n = 5) (χ^2^ = 1.543, p = 0.2142), and in the protein group (χ^2^ = 2.100, p = 0.1473), it increased from 90% (n = 19) to 100% (n = 21).

## DISCUSSION AND CONCLUSION

The results of this study suggest that eHealth reduced IBS symptom severity and improved quality of life. There are three main reasons for the reduction in symptoms measured by the IBS-SI as the primary outcome, and the improvement in quality of life measured by the IBS-QOL as a secondary outcome. The first reason was that the eHealth program had extensive food-related content. The second reason was that the eHealth program had a wide range of non-food content in a variety of genres. A third was the accessibility of the eHealth program.

First, several sections of the eHealth program included information on food sections such as “diet and the digestive system” and “dietary management,” and within each section was specific and detailed content related to IBS symptoms. Existing IBS eHealth focuses primarily on diets such as FODMAPs [34, 35, 36, 37] and probiotics[34, 35], and it is already known that these approaches contribute to the improvement of IBS symptoms. In other words, learning about food in this study may also have normalized food intake. The secondary outcome of this study, food intake, were normalizes by the eHealth program. Improvement of food intake normalized the intestinal environment. For example, a previous study comparing the impact of a low FODMAP diet on IBS symptoms found an improvement in intestinal health alongside improvement in IBS symptoms [27]. It is also possible that the improvement in food intake through learning about food by using eHealth program in our study, may have reduced Cyanobacterial occupancy. In a study of pediatric IBS, Cyanobacteria were found to be rich in those with high fructan sensitivity [44]. In an animal study, diarrhea model mice had significantly higher levels of Cyanobacteria than normal mice, indicating that Cyanobacteria are reduced Chinese herbal prescriptions [45]. These facts suggest that Cyanobacteria are involved in the exacerbation of symptoms of diarrheal IBS. In this study, participants had more diarrheal (12 IBS-D and 12 IBS-M, 60%) in both the eHealth and TAU groups.

Elaborating on the second reason, the wide variety of content in the eHealth program (Table 1) may have contributed to the reduction of IBS-SI and improved quality of life. The original self-help guidebook contained evidence-based information and techniques associated with IBS symptom improvement [31, 32], and similar content was included in the eHealth version used in this study. In addition, the eHealth program offered a wide range of content on non-dietary measure, such as exercise [25], cognitive-behavioral therapy[20], and relaxation[16] known to contribute to IBS symptom reduction.

As a third reason, the ease of access to eHealth programs may have contributed to the reduction of IBS symptoms and improved quality of life. The eHealth program created in this study [38] was designed to be accessible not only through computers but also tablets or smart phones. Such information and communication technologies facilitated access to and the use of medical and health information, thereby assisting in self-management [46]. Additionally, the system design enabled content to be downloaded and stored locally on personal devices. Hence, content could be accessed whenever and wherever the patient wished to learn. These characteristics of the eHealth program may have contributed to the reduction of IBS symptoms and improved quality of life.

The strength of this study is its observation of extremely good compliance. Because the eHealth program was designed to be accessed whenever and wherever, all participants accessed all content at least once. Therefore, all the collected data could be used for the statistical analyses as there were no dropouts.

The first limitation of this study is that the participants’ diagnosis of IBS was based on a self-reported questionnaire rather than a physician’s assessment. Therefore, IBS may have been overestimated or inappropriately diagnosed in some participants. The second limitation is that the study did not adjust for food intake as a confounding factor. The assessment of food intake in this study was qualitative only, and dietary changes strongly affect IBS symptoms [34, 35, 36, 37]. In the future, a reliable quantitative evaluation is needed to assess the potential impact of the eHealth program used in this study on changes in food intake. The third limitation is that it does not adjust for lifestyle factors other than diet, such as sleep [47] and exercise [25], which also affect IBS symptoms, although to a lesser extent. In future, it is necessary to evaluate the strength of the influence of individual lifestyle factors, such as sleep and exercise, in addition to diet, on IBS symptoms and quality of life.

In conclusion, the implementation of eHealth for IBS was shown to reduce IBS symptoms.

## Data Availability

All data relevant to the study are included in the article or uploaded as supplementary information.

## ACKNOWLEDGEMENTS

The authors would like to thank the participants of this study. This study was supported by Waseda University, Saitama Prefectural University, and Nagasaki University. Dr. Anne Kennedy of Southampton University, who passed away before this paper was completed, strongly encouraged us to pursue this research. We would like to thank her, and her family, friends, and colleagues.

## REFERENCES

1 Mayer EA, Raybould HE. Role of visceral afferent mechanisms in functional bowel disorders. Gastroenterology 1990;99:1688–704.

2 Drossman DA, Sandler RS, McKee DC, et al., Bowel patterns among subjects not seeking health care. Use of a questionnaire to identify a population with bowel dysfunction. Gastroenterology 1982;83:529–34.

3 Ritchie J. Pain from distension of the pelvic colon by inflating a balloon in the irritable colon syndrome. Gut 1973;14:125–32.

4 Tayama J, Nakaya N, Hamaguchi T, et al. Effects of personality traits on the manifestations of irritable bowel syndrome. Biopsychosoc Med 2012;6:20.

5 Sperber AD, Dumitrascu D, Fukudo S, et al. The global prevalence of IBS in adults remains elusive due to the heterogeneity of studies: a Rome Foundation working team literature review. Gut 2017;66:1075–82.

6 Gralnek IM, Hays RD, Kilbourne A, et al. The impact of irritable bowel syndrome on health-related quality of life. Gastroenterology 2000;119:654–60.

7 Kanazawa M, Drossman DA, Shinozaki M, et al., Translation and validation of a Japanese version of the irritable bowel syndrome-quality of life measure (IBS-QOL-J). Biopsychosoc Med 2007;1:6.

8 Jerndal P, Ringstrom G, Agerforz P, et al. Gastrointestinal-specific anxiety: an important factor for severity of GI symptoms and quality of life in IBS. Neurogastroenterol Motil 2010;22:646–e179.

9 Jamali R, Jamali A, Poorrahnama M, et al. Evaluation of health-related quality of life in irritable bowel syndrome patients. Health Qual Life Outcomes 2012;10:12.

10 Maxion-Bergemann S, Thielecke F, Abel F, et al. Costs of irritable bowel syndrome in the UK and US. Pharmacoeconomics 2006;24:21–37.

11 Lacy BE, Pimentel M, Brenner DM, et al. ACG clinical guideline: Management of irritable bowel syndrome. Am J Gastroenterol 2021;116:17–44.

12 Fukudo S, Okumura T, Inamori M, et al. Evidence-based clinical practice guidelines for irritable bowel syndrome 2020. J Gastroenterol 2021;56:193–217.

13 Sagami Y, Shimada Y, Tayama J, et al. Effect of a corticotropin releasing hormone receptor antagonist on colonic sensory and motor function in patients with irritable bowel syndrome. Gut 2004;53:958–64.

14 Longstreth GF, Thompson WG, Chey WD, et al. Functional bowel disorders. Gastroenterology 2006;130:1480–91.

15 Saito K, Kanazawa M, Fukudo S. Colorectal distention induces hippocampal noradrenaline release in rats: an in vivo microdialysis study. Brain Res 2002;947:146–9.

16 Blanchard EB, Greene B, Scharff L, et al. Relaxation training as a treatment for irritable bowel syndrome. Biofeedback Self Regul 1993;18:125–32.

17 Whorwell PJ, Prior A, Colgan SM. Hypnotherapy in severe irritable bowel syndrome: further experience. Gut 1987;28:423–5.

18 Roberts L, Wilson S, Singh S, et al. Gut-directed hypnotherapy for irritable bowel syndrome: piloting a primary care-based randomised controlled trial. Br J Gen Pract 2006;56:115–21.

19 Lackner JM, Jaccard J, Keefer L, et al. Improvement in gastrointestinal symptoms after cognitive behavior therapy for refractory irritable bowel syndrome. Gastroenterology 2018;155:47–57.

20 Sugaya N, Shirotsuki K, Nakao M. Cognitive behavioral treatment for irritable bowel syndrome: a recent literature review. Biopsychosoc Med 2021;15:23.

21 Naliboff BD, Smith SR, Serpa JG, et al. Mindfulness-based stress reduction improves irritable bowel syndrome (IBS) symptoms via specific aspects of mindfulness. Neurogastroenterol Motil 2020;32:e13828.

22 Henrich JF, Gjelsvik B, Surawy C, et al. A randomized clinical trial of mindfulness-based cognitive therapy for women with irritable bowel syndrome-Effects and mechanisms. J Consult Clin Psychol 2020;88:295–310.

23 Labus J, Gupta A, Gill HK, et al. Randomised clinical trial: symptoms of the irritable bowel syndrome are improved by a psycho-education group intervention. Aliment Pharmacol Ther 2013;37:304–15.

24 Ringstrom G, Storsrud S, Lundqvist S, et al. Development of an educational intervention for patients with Irritable Bowel Syndrome (IBS): a pilot study. BMC Gastroenterol 2009;9:10.

25 Hamaguchi T, Tayama J, Suzuki M, et al. The effects of locomotor activity on gastrointestinal symptoms of irritable bowel syndrome among younger people: An observational study. PLoS One 2020;15:e0234089.

26 Johannesson E, Simren M, Strid H, et al. Physical activity improves symptoms in irritable bowel syndrome: a randomized controlled trial. Am J Gastroenterol 2011;106:915–22.

27 McIntosh K, Reed DE, Schneider T, et al. FODMAPs alter symptoms and the metabolome of patients with IBS: a randomised controlled trial. Gut 2017;66:1241–51.

28 Staudacher HM, Whelan K. The low FODMAP diet: recent advances in understanding its mechanisms and efficacy in IBS. Gut 2017;66:1517–27.

29 O’Sullivan MA, Mahmud N, Kelleher DP, et al. Patient knowledge and educational needs in irritable bowel syndrome. Eur J Gastroenterol Hepatol 2000;12:39–43.

30 Robinson A, Lee V, Kennedy A, Middleton L, et al. A randomised controlled trial of self-help interventions in patients with a primary care diagnosis of irritable bowel syndrome. Gut 2006;55:643–8.

31 Kennedy A, Robinson A, Rogers A. Incorporating patients’ views and experiences of life with IBS in the development of an evidence based self-help guidebook. Patient Educ Couns 2003;50:303–10.

32 Schneider A, Rosenberger S, Bobardt J, et al. Self-help guidebook improved quality of life for patients with irritable bowel syndrome. PLoS One 2017;12:e0181764.

33 Ankersen DV, Carlsen K, Marker D, et al. Using eHealth strategies in delivering dietary and other therapies in patients with irritable bowel syndrome and inflammatory bowel disease. J Gastroenterol Hepatol 2017;32 Suppl 1:27–31.

34 Ankersen DV, Weimers P, Bennedsen M, et al. Long-term effects of a web-based low-FODMAP diet versus probiotic treatment for irritable bowel syndrome, including shotgun analyses of microbiota: Randomized, double-crossover clinical trial. J Med Internet Res 2021;23:e30291.

35 Pedersen N, Andersen NN, Vegh Z, et al. Ehealth: low FODMAP diet vs Lactobacillus rhamnosus GG in irritable bowel syndrome. World J Gastroenterol 2014;20:16215–26.

36 Pedersen N, Vegh Z, Burisch J, et al. Ehealth monitoring in irritable bowel syndrome patients treated with low fermentable oligo-, di-, mono-saccharides and polyols diet. World J Gastroenterol 2014;20:6680–4.

37 Pedersen N. EHealth: self-management in inflammatory bowel disease and in irritable bowel syndrome using novel constant-care web applications. EHealth by constant-care in IBD and IBS. Dan Med J 2015;62:B5168.

38 Hori M, Ono S, Yamaji K, et al. A suitable m-learning system using e-book for developing countries. In CSEDU 2016;2:408–15.

39 Shinozaki M, Kanazawa M, Sagami Y, et al. Validation of the Japanese version of the Rome II modular questionnaire and irritable bowel syndrome severity index. J Gastroenterol 2006;41:491–4.

40 Francis CY, Morris J, Whorwell PJ. The irritable bowel severity scoring system: a simple method of monitoring irritable bowel syndrome and its progress. Aliment Pharmacol Ther 1997;11:395–402.

41 Patrick DL, Drossman DA, Frederick IO, DiCesare J, Puder KL. Quality of life in persons with irritable bowel syndrome: development and validation of a new measure. Dig Dis Sci 1998;43:400–11.

42 Bolyen E, Rideout JR, Dillon MR, et al. Reproducible, interactive, scalable and extensible microbiome data science using QIIME 2. Nat Biotechnol 2019;37:852–7.

43 Segata N, Izard J, Waldron L, et al. Metagenomic biomarker discovery and explanation. Genome Biol 2011;12:R60.

44 Chumpitazi BP, Hoffman KL, Smith DP, et al. Fructan-sensitive children with irritable bowel syndrome have distinct gut microbiome signatures. Aliment Pharmacol Ther 2021;53:499–509.

45 Li L, Cui H, Li T, et al. Synergistic effect of berberine-based Chinese medicine assembled nanostructures on diarrhea-predominant irritable bowel syndrome in vivo. Front Pharmacol 2020;11:1210.

46 Ossebaard HC, Van Gemert-Pijnen L. eHealth and quality in health care: implementation time. Int J Qual Health Care 2016;28:415–9.

47 Lee SK, Yoon DW, Lee S, et al. The association between irritable bowel syndrome and the coexistence of depression and insomnia. J Psychosom Res 2017;93:1–5.

